# Tumor CTR1 and serum copper dynamics reveal a coordinated copper axis linked to high-grade triple-negative breast cancer biology

**DOI:** 10.64898/2025.12.18.25342516

**Authors:** Vinit C. Shanbhag, Nikita Gudekar, Muhammad Yasir, Kristyn Conrad, Samuel Anakpeba-Dinguyella, Parshad Sutar, Praveen Rao, Michael Petris, Linda Vahdat, Christos Papageorgiou

## Abstract

**Background:** Copper is an essential nutrient required for energy production, antioxidant defense, and connective tissue maturation, yet has emerged as a metabolic vulnerability in cancer. CTR1 (*SLC31A1*), the high-affinity copper importer, mediates cellular copper uptake, and its upregulation may signal increased copper demand in tumor cells. The dynamics of copper regulation across tumor growth, aggressiveness, and treatment resistance remain poorly defined in breast cancer. We investigated whether CTR1 expression and systemic copper changes reflect a coordinated tumor-systemic copper axis

**Methods:** A retrospective dataset of 1632 breast cancer patients receiving neoadjuvant chemotherapy was analyzed to compare CTR1 gene expression between responders and non-responders across molecular subtypes and tumor grades. Findings were extended to a prospective neoadjuvant cohort in which paired pre-and post-treatment serum copper levels were measured. ΔCopper (post–pre change) was correlated with subtype, grade, response, and tumor size

**Results:** CTR1 expression was significantly higher in triple-negative breast cancer (TNBC) non-responders than responders (*P* = 0.0021), particularly in grade 3 tumors (*P* = 0.0035), with no difference in luminal subtypes. In the prospective cohort, △Copper was positive predominantly in TNBC and strongly grade-dependent: all grade 3 TNBCs exhibited copper elevation post-therapy, whereas all grade 2 TNBCs showed negative △Copper (*P* = 0.034). The only relapse in the cohort, a TNBC non-responder, exhibited persistently positive △Copper at follow-up and relapse, whereas non-responders from other subtypes showed near-zero or negative △Copper (*P* = 0.011). Baseline serum copper was higher in patients with smaller (clinical T1) versus larger (T2–T3) tumors (*P* = 0.033)

**Conclusions:** Parallel CTR1 upregulation in tumors and systemic copper elevation post-therapy suggest a coordinated copper mobilization program in high-grade TNBC. These integrated retrospective and prospective findings link copper transport to therapy response and tumor aggressiveness, highlighting copper biology as a potential therapeutic axis in breast cancer.

## 1. Background

Breast cancer remains a leading cause of cancer-related morbidity and mortality worldwide, with heterogeneous outcomes across molecular subtypes. Despite advances in targeted therapy and immunotherapy, there is an unmet need for biomarkers that can both capture disease biology in real time and inform treatment decisions.

Copper (Cu) is an essential nutrient required for energy production, antioxidant defense, and connective tissue maturation, yet its metabolism has emerged as a unique vulnerability in cancer.^1^ Cellular copper uptake is primarily mediated by the high-affinity transporter CTR1 (*SLC31A1*), the principal gateway for copper entry and a key regulator of intracellular copper availability.^2, 3^ Global Ctr1 deletion in mice causes embryonic lethality,^4^ and intestinal loss results in systemic copper deficiency,^5^ underscoring its physiological importance. CTR1-driven copper uptake supports several oncogenic programs. Copper-dependent activation of the MAPK–ERK pathway and ULK1/2 signaling links CTR1-mediated copper transport to cell growth, autophagy, and tumor progression.^6, 7^

Elevated serum copper has been associated with adverse prognosis, and intracellular copper availability supports multiple tumor-promoting pathways.^1^ These include angiogenesis through VEGF signaling,^8^ mitochondrial respiration via cytochrome c oxidase,^9, 10^ antioxidant defense through superoxide dismutase,^11, 12^ extracellular matrix remodeling through lysyl oxidases,^13–15^ and regulation of signaling kinases such as MEK1 and ULK1.^6, 7^ In an effort to counteract these oncogenic effects of copper, a phase II clinical study of systemic copper depletion with tetrathiomolybdate (TTM) in high-risk breast cancer patients demonstrated feasibility and suggested benefit in triple-negative breast cancer (TNBC).^16^

Despite these insights, how copper mobilization and CTR1-mediated uptake are regulated across tumor grades, aggressiveness, and treatment contexts remains poorly understood. Most prior studies have examined static baseline copper levels rather than dynamic changes in copper transport or systemic copper availability. To address this gap, we conducted a large retrospective analysis assessing post-chemotherapy CTR1 expression in responders versus non-responders across breast cancer subtypes and tumor grades, complemented by a prospective exploratory study measuring paired pre-and post-treatment serum copper levels (△Copper) in the neoadjuvant setting. This integrative approach enables parallel evaluation of tumor copper demand and systemic copper mobilization, providing insight into a coordinated tumor-systemic copper axis in breast cancer.

## 2. Methods

### 2.1 Study design

This study incorporated a retrospective gene-expression analysis and a prospective observational cohort to examine copper biology in breast cancer (Figure 1). The retrospective arm evaluated *SLC31A1* (CTR1) expression in public datasets, while the prospective arm measured serum copper dynamics (△Copper) during neoadjuvant therapy. Both datasets were analyzed independently and then compared to identify convergent trends between tissue gene expression and systemic copper changes.

**Figure 1.**
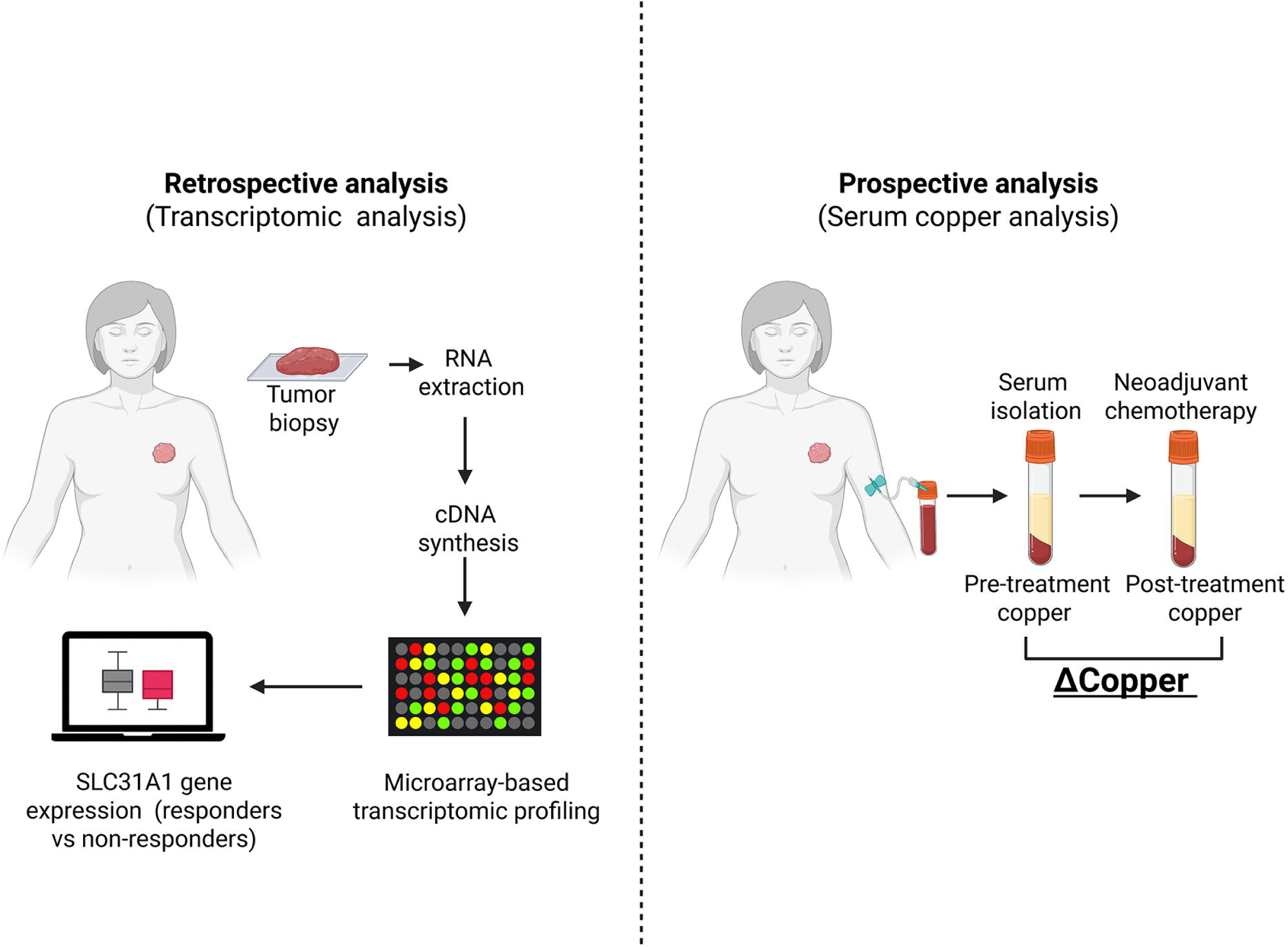
Overview of retrospective and prospective study design. The retrospective arm compared tumor *SLC31A1* expression in responders and non-responders after neoadjuvant therapy, and the prospective arm measured serum copper changes (△Copper) before and after neoadjuvant therapy. Abbreviations: △, delta.

### 2.2 Retrospective analysis of *SLC31A1* gene expression

Expression of *SLC31A1* (CTR1) was analyzed using the ROCplot.org platform,^17^ which integrates transcriptomic and treatment-response data from 36 publicly available breast-cancer datasets. The analysis focused on patients with pathologic response data (n = 1,632) following neoadjuvant chemotherapy. Subgroup analyses were performed by molecular subtype (triple-negative, Luminal A, Luminal B) and by tumor grade within the TNBC cohort. Differences in *SLC31A1* expression between responders and non-responders were assessed using the Mann–Whitney U test, and predictive performance summarized by the area under the receiver-operating-characteristic curve (AUC). In biomarker assessment, AUC values near 0.5 indicate no predictive value; AUC <0.6 reflects limited utility; AUC 0.6–0.7 represents a good biomarker with potential clinical relevance; and AUC >0.7 denotes a high-quality candidate with strong discriminatory performance.

### 2.3 Subtyping and clinical classification

Breast cancer subtypes were defined by estrogen receptor (ER), progesterone receptor (PR), and human epidermal growth factor receptor 2 (HER2) status assessed by immunohistochemistry and confirmed by fluorescence in situ hybridization (FISH). Triple negative breast cancer (TNBC) was defined as ER <1%, PR <1%, and HER2 negative by IHC or FISH. Tumor grade and pathologic stage were from final pathology reports. Treatment response was classified by the treating clinician as responder or non-responder.

### 2.4 Serum collection and copper measurement

Since serum copper is not materially affected by time of day or postprandial interval,^18, 19^ patients were not required to have fasted prior to blood draws, thus minimizing disruption of standard clinical workflow in the neoadjuvant setting.^20^ Peripheral blood was collected by venipuncture at three time points: diagnosis (before therapy), completion of neoadjuvant chemotherapy, and biopsy-confirmed relapse. Serum was separated and stored at −80°C. Total copper was quantified by ICP-MS (Agilent 8900 ICP-QQQ) using standard trace-metal workflows.

### 2.5 Clinical variables captured

Clinical variables were collected as part of routine clinical care and research protocols and are reported in de-identified form.

### 2.6 Primary exposure

△Copper (post-pre treatment serum copper, µg/kg) represented the primary exposure. A positive value reflects higher copper after therapy. The first post-therapy sample was used for analysis; later follow-up samples, including the relapse case, were summarized descriptively.

### 2.7 Statistical analysis (prospective cohort)

Data are summarized as medians with interquartile ranges (IQRs); means, minima, and maxima are reported where informative. Associations between △Copper and clinicopathologic variables were examined using nonparametric methods. Correlations were evaluated with Spearman’s rank (ρ) and two-sided P values. Group comparisons used Mann–Whitney U tests; comparisons across multiple subtypes used Kruskal–Wallis tests. Analyses were performed in Python (v3.x) with pandas library for data handling, NumPy for numerical computation, and SciPy (scipy.stats) for statistical testing. Figures and graphs were generated in GraphPad Prism (v10).

## 3. Results

### 3.1 Patient characteristics

A total of 21 patients with breast cancer were included in the prospective cohort. Demographic characteristics are summarized in Table 1 and are presented in de-identified form in accordance with preprint data-sharing requirements. Clinical variables were evaluated in relation to serum copper dynamics, tumor subtype, grade, and treatment response and are analyzed in subsequent sections.

**Table 1.** Demographic characteristics of breast cancer patients (N = 21)

| **Patient ID** | **Age range**<br>**(years)** | **Sex** |
| --- | --- | --- |
| BCP-001 | 60–64 | F |
| BCP-002 | 60–64 | F |
| BCP-003 | 35–39 | F |
| BCP-004 | 25–29 | F |
| BCP-005 | 60–64 | F |
| BCP-006 | 35–39 | F |
| BCP-007 | 50–54 | F |
| BCP-008 | 50–54 | F |
| BCP-009 | 35–39 | F |
| BCP-010 | 70–74 | F |
| BCP-011 | 40–44 | F |
| BCP-012 | 60–64 | F |
| BCP-013 | 35–39 | F |
| BCP-014 | 55–59 | F |
| BCP-015 | 70–74 | F |
| BCP-016 | 70–74 | F |
| BCP-017 | 55–59 | F |
| BCP-018 | 60–64 | F |
| BCP-019 | 50–54 | F |
| BCP-020 | 45–49 | F |
| BCP-021 | 70–74 | M |

Ages are presented as non-overlapping 5 year ranges to minimize the risk of participant reidentification. Patient IDs shown are anonymized for presentation and do not correspond to original identifiers.

### 3.2 Retrospective analysis of *SLC31A1* expression and pathologic response

The integrated study design is shown in Figure 1. To evaluate whether copper import relates to chemotherapy response, *SLC31A1* expression was analyzed in 1,632 breast-cancer patients with pathologic response following neoadjuvant chemotherapy.^17^ Figure 2A illustrates CTR1 localization on tumor cells and its role in facilitating copper uptake.

**Figure 2.**
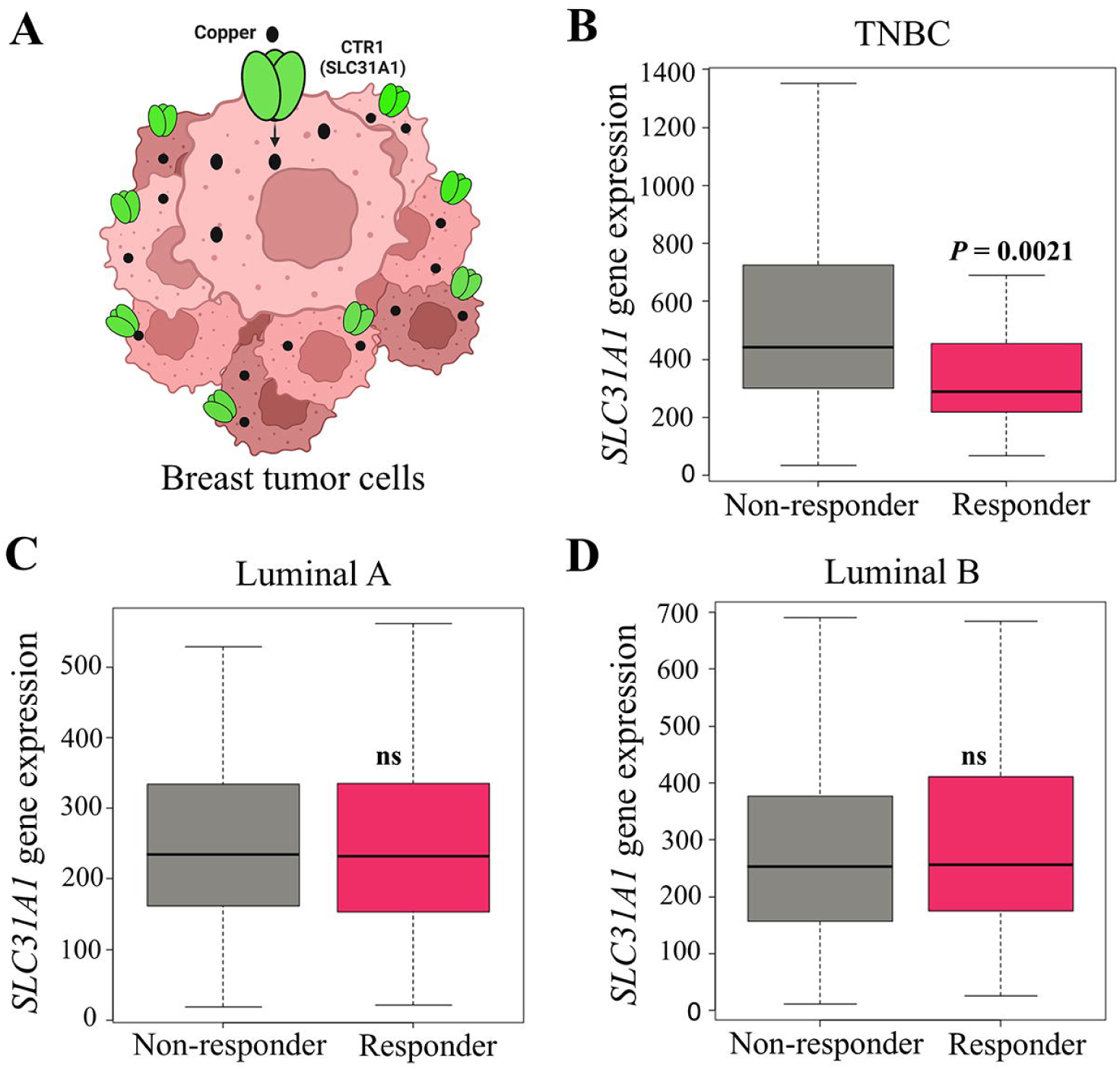
CTR1 function and *SLC31A1* expression by pathologic response. (A) Illustration of CTR1 on the tumor cell surface mediating copper uptake. (B–D) *SLC31A1* transcript levels in responders versus non-responders across breast cancer subtypes. Abbreviations: ns, not significant; TNBC, triple-negative breast cancer.

Across molecular subtypes, *SLC31A1* expression was highest in triple-negative breast cancer (TNBC) and significantly elevated in non-responders (n = 277) compared with responders (*n* = 196, *P* = 0.0021; Figure 2B; Table 2). The corresponding AUC was 0.682, indicating good predictive potential for distinguishing TNBC non-responders. No significant differences were observed in Luminal A (n = 341 vs 134, *P* = 0.44, AUC = 0.509; Figure 2C) or Luminal B (n = 372 vs 119, *P* = 0.36, AUC = 0.522; Figure 2D; Table 2). Within TNBC, *SLC31A1* expression also varied by histologic grade. In grade 3 tumors, non-responders (*n* = 172) showed significantly higher *SLC31A1* expression than responders (*n* = 89, *P* = 0.0035; Figure 3A; Table 3), with an AUC of 0.717, reflecting stronger discriminatory ability in high-grade disease. In contrast, grade 2 tumors showed no significant difference between non-responders and responders (*n* = 42 vs 13, *P* = 0.54, AUC = 0.576; Figure 3B; Table 3).

**Figure 3.**
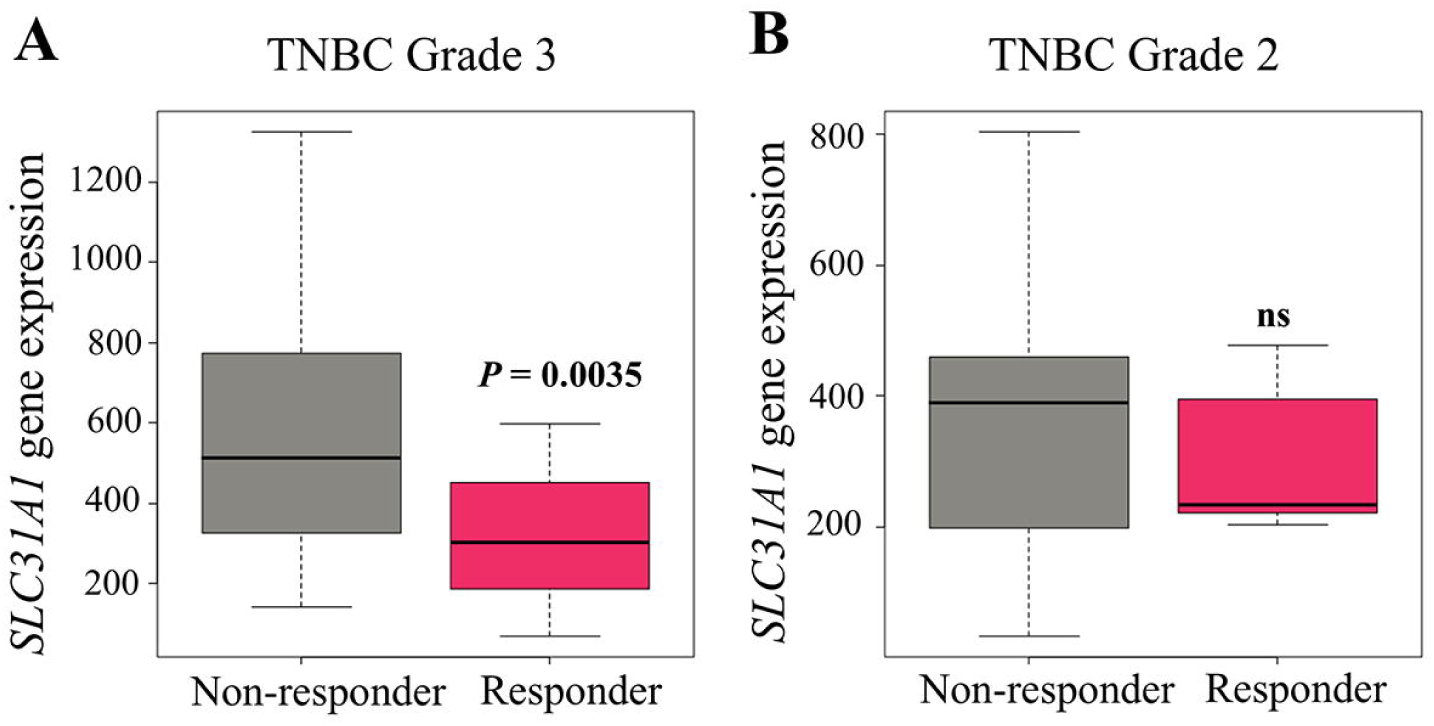
*SLC31A1* expression in TNBC by tumor grade and response. Box plots show *SLC31A1* expression in responders and non-responders with grade 3 (A) or grade 2 (B) TNBC. Abbreviations: ns, not significant; TNBC, triple-negative breast cancer.

**Table 2.** Sample distribution and statistical comparison for *SLC31A1* analysis across breast cancer subtypes.

| Subtype | Non-responders (n) | Responders (n) | P value (Mann Whitney U) |
| --- | --- | --- | --- |
| TNBC | 277 | 196 | 0.0021 |
| Luminal A | 341 | 134 | Not significant |
| Luminal B | 372 | 119 | Not significant |

Abbreviations: TNBC, triple-negative breast cancer.

**Table 3.** Sample distribution and statistical comparison for *SLC31A1* analysis by tumor grade in triple-negative breast cancer.

| **Subtype/**<br>**Grade** | **Non-responders (n)** | **Responders (n)** | **P value (Mann Whitney U)** |
| --- | --- | --- | --- |
| TNBC grade 3 | 172 | 89 | 0.0035 |
| TNBC grade 2 | 42 | 13 | 0.54 (not significant) |

Abbreviations: TNBC, triple-negative breast cancer.

These findings indicate that *SLC31A1* upregulation associates with chemotherapy nonresponse and aggressive tumor grade specifically in TNBC and provided the rationale for evaluating dynamic systemic copper changes in the prospective cohort.

### 3.3 Distribution of △Copper across breast cancer molecular subtypes

To explore the dynamic behavior of serum copper following treatment, delta copper (△Copper), defined as the change in serum copper concentration from pre-to post-treatment, was quantified across breast cancer molecular subtypes, representing a novel and dynamic metric introduced in this cohort. Median △Copper values differed by subtype, with TNBC (n=6) showing the highest median increase of 123.0 µg/kg (interquartile range [IQR], 433.8 µg/kg), followed by HR+/HER2− (n=6) at 98.5 µg/kg (IQR, 263.3 µg/kg), whereas HR+/HER2+ (n=6) and HR−/HER2+ (n=3) exhibited median decreases of –89.5 µg/kg (IQR, 407.5 µg/kg) and –119.0 µg/kg (IQR, 225.5 µg/kg), respectively (Figure 4; Table 4). Although overall distributions did not differ significantly among subtypes (Kruskal–Wallis H = 1.71, *P* = 0.63), TNBC displayed the broadest range of values (–416 to +978 µg/kg) and a trend toward higher △Copper compared with other groups.

**Figure 4.**
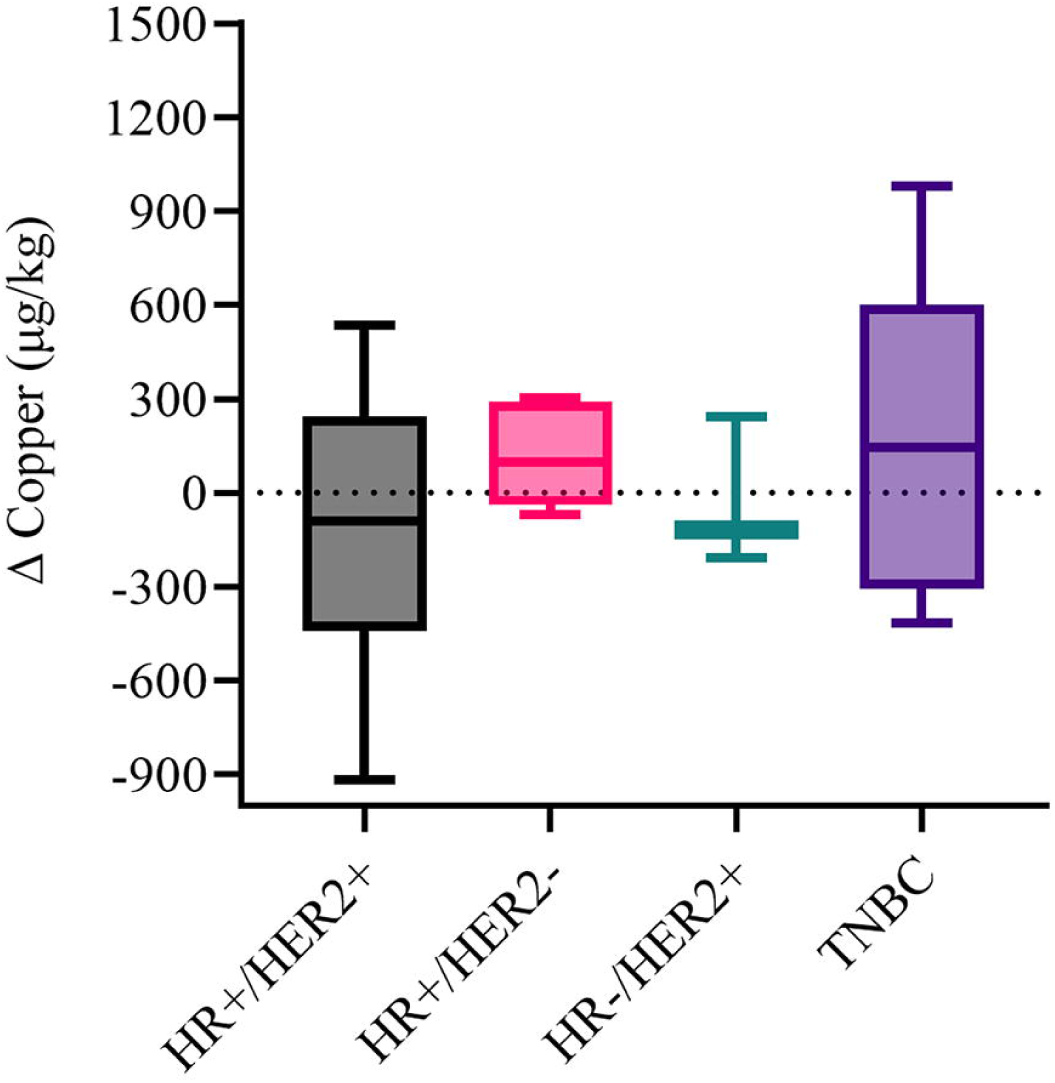
△Copper levels in breast cancer patients by molecular subtype. Box plots show the distribution of △Copper values (posttreatment − pretreatment) across subtypes. The horizontal line within each box represents the median; box edges represent the interquartile range (IQR); whiskers indicate the minimum and maximum values. Abbreviations: △, delta; IQR, interquartile range; TNBC, triple-negative breast cancer.

**Table 4.** △Copper levels in breast cancer patients by molecular subtype (N=21)

| **Subtype** | **n** | **Median**<br>**ΔCu μg/kg** | **Mean**<br>**ΔCu μg/kg** | **IQR** | **Min** | **Max** |
| --- | --- | --- | --- | --- | --- | --- |
| HR+/HER2+ | 6 | -89.5 | -116.8 | 407.5 | -919 | 534 |
| HR+/HER2- | 6 | 98.5 | 114.5 | 263.3 | -72 | 304 |
| HR-/HER2+ | 3 | -119.0 | -28.7 | 225.5 | -209 | 242 |
| TNBC | 6 | 123.0 | 126.2 | 433.8 | -416 | 978 |
| Total | 21 |  |  |  |  |  |

Abbreviations: Cu, copper; HER2, human epidermal growth factor receptor 2; HR, hormone receptor; IQR, interquartile range; TNBC, triple-negative breast cancer.

### 3.4 Distinct △Copper pattern in the TNBC relapse case

Because treatment resistance may be associated with distinct copper dynamics, △Copper values were examined in non-responders across subtypes (Figure 5). In HR+/HER2+ (n=4) and HR+/HER2− (n=5) subtypes, most non-responders showed near-zero or negative △Copper, whereas in TNBC, the single non-responder, who subsequently relapsed, demonstrated consistently positive △Copper values at both follow-up timepoints, which were specifically measured to track changes post-treatment and at cancer relapse (Figure 5A). This difference among non-responders reached statistical significance (Spearman ρ = 0.73; *P* = 0.011). In Figure 5B, longitudinal serum copper trajectories for this relapse case showed progressive increases at each timepoint: from 932 µg/kg pre-treatment to 1,188 µg/kg at first follow-up and 1,533 µg/kg at cancer relapse.

**Figure 5.**
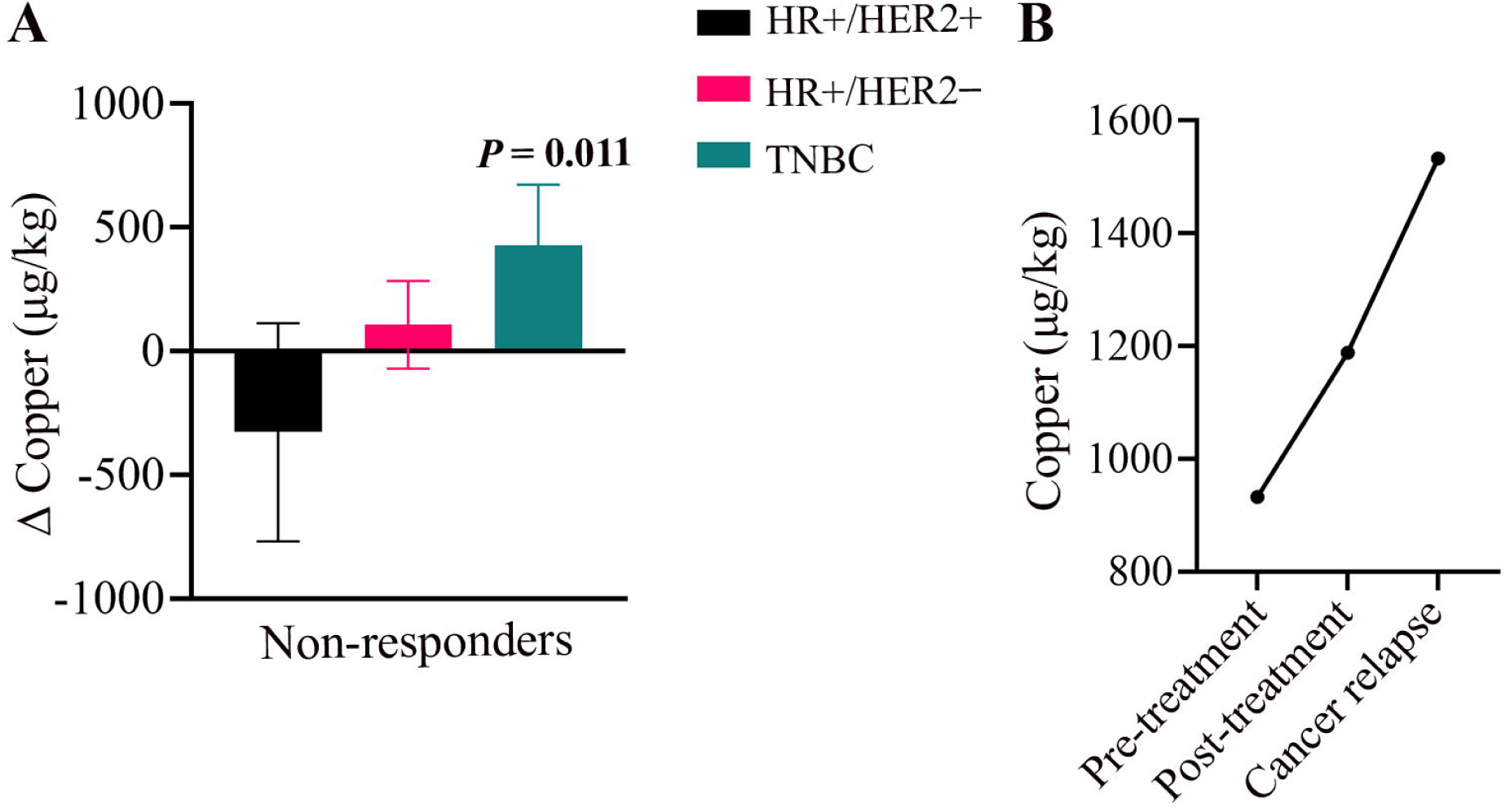
△Copper and relapse in breast cancer non-responders. (A) Distribution of △Copper values in non-responders across molecular subtypes. (B) Serum copper trajectory of the single TNBC patient who relapsed, showing changes from pretreatment to posttreatment and relapse follow-up. Abbreviations: △, delta; TNBC, triple-negative breast cancer.

### 3.5 Tumor grade correlates with △Copper in TNBC

We next assessed △Copper by tumor grade within each subtype, a recognized marker for tumor aggressiveness (Figure 6). In the TNBC group, a striking pattern emerged: all the patients harboring grade 3 tumors (n=4) exhibited positive △Copper values, whereas all the patients harboring grade 2 tumors (n=2) displayed negative △Copper values (Figure 6A). This association was statistically significant (Spearman ρ = 0.79; *P* = 0.034). Conversely, no consistent or significant correlations were identified between tumor grade and △Copper in the HR+/HER2+, HR+/HER2−, or HR−/HER2+ subtypes (Figures 6B-D). These findings highlight a grade-related copper dynamic unique to TNBC in this cohort.

**Figure 6.**
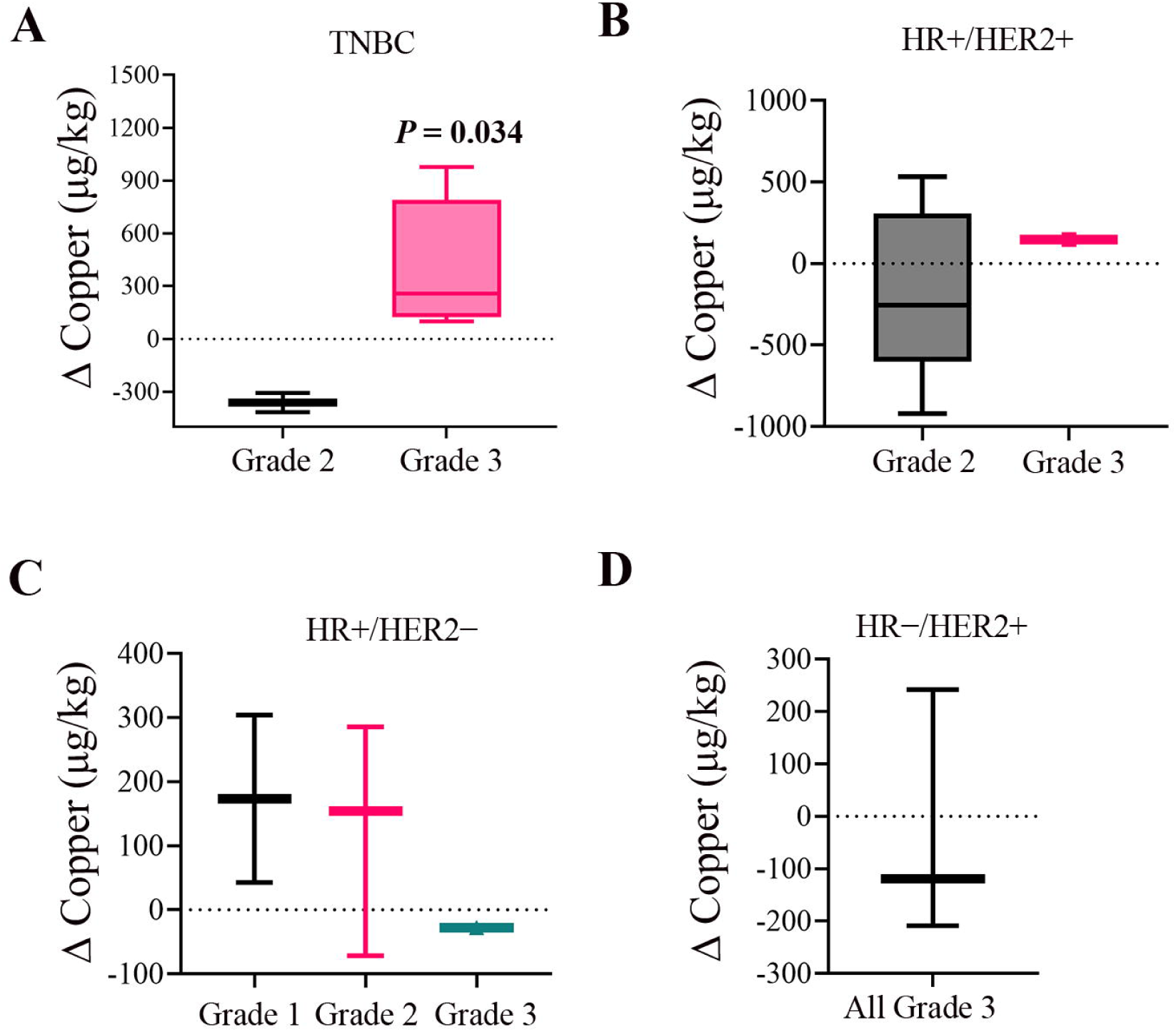
△Copper levels stratified by tumor grade within molecular subtypes. Box plots display △Copper values (posttreatment − pretreatment) by tumor grade within each molecular subtype: (A) TNBC, (B) HR+/HER2+, (C) HR+/HER2−, and (D) HR−/HER2+. The horizontal line within each box represents the median; box edges indicate the interquartile range (IQR); and whiskers extend to the minimum and maximum values. Abbreviations: △, delta; HR, hormone receptor; IQR, interquartile range; TNBC, triple-negative breast cancer.

### 3.6 Inverse association between tumor size and pretreatment serum copper

We next examined whether pretreatment serum copper levels were associated with clinical tumor size at diagnosis (Figure 7). Prior preclinical studies have reported higher intratumoral copper concentrations in smaller tumors,^10^ and thus we evaluated whether this relationship extended to systemic serum copper. Female patients were grouped by clinical tumor stage as having T1 tumors (≤2 cm; n = 3) or T2–T3 tumors (>2 cm; n = 16). Median pretreatment serum copper levels were significantly higher in patients with smaller T1 tumors compared with those harboring larger T2–T3 tumors (1,435 µg/kg [IQR, 166 µg/kg] vs 1,015 µg/kg [IQR, 268 µg/kg]; Mann-Whitney U = 43, *P* = 0.033).

**Figure 7.**
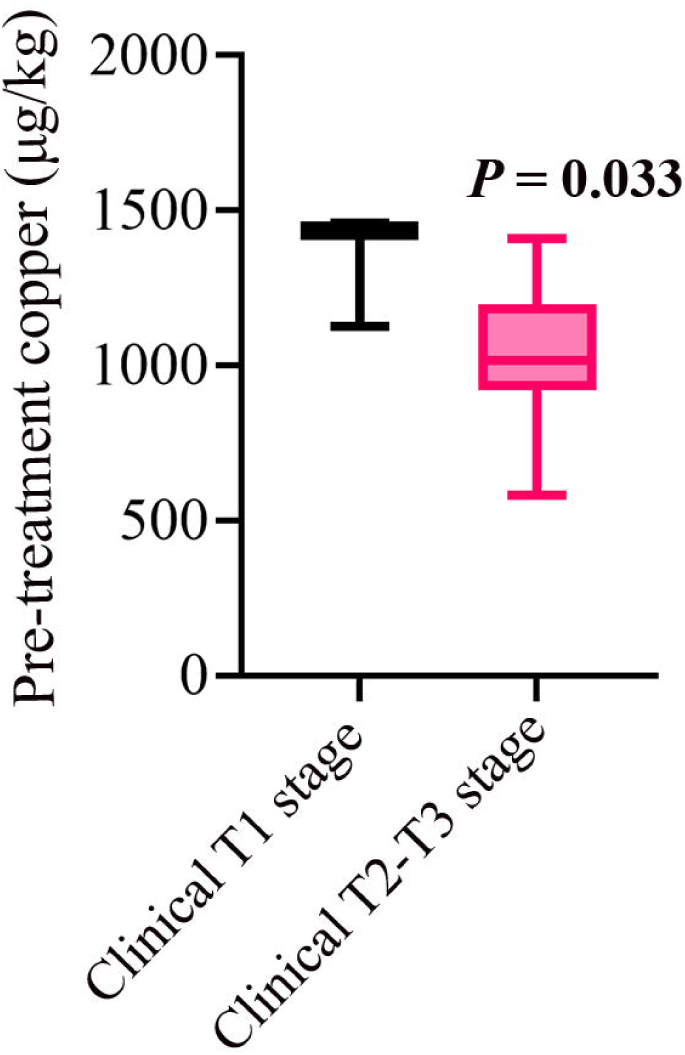
Serum copper levels by clinical T stage. Box plots show serum copper concentrations (µg/kg) for patients with stage T1 (n = 3) and stage T2–T3 (n = 16) breast tumors. The horizontal line within each box indicates the median; box edges represent the interquartile range (IQR); and whiskers denote the minimum and maximum values. Abbreviations: IQR, interquartile range.

## 4. Discussion

Copper supports multiple metabolic and signaling processes essential to cancer progression, including mitochondrial respiration, redox maintenance, angiogenesis, and extracellular matrix (ECM) remodeling.^1^ Clinical studies of systemic copper chelation in high-risk triple-negative breast cancer (TNBC) suggest that copper metabolism can be therapeutically targeted in this subtype.^16^ However, key questions remain: Why does TNBC show such a strong association with copper biology? Is copper linked to tumor aggressiveness and resistance to neoadjuvant chemotherapy? And can copper-associated biomarkers guide personalized treatment or copper-chelation strategies?

To begin addressing these questions, we conducted the first integrated analysis combining paired serum copper measurements in a prospective neoadjuvant setting with retrospective tumor CTR1 copper importer gene expression linked to neoadjuvant chemotherapy response. This allowed us to assess copper uptake at the tumor level (CTR1) alongside dynamic systemic copper changes (△Copper) during treatment, an aspect of copper biology not previously evaluated in breast cancer patients.

A consistent and clinically meaningful pattern emerged across both datasets: high-grade TNBC engages a coordinated copper axis. In the retrospective analysis of 1,632 patients, CTR1 (*SLC31A1*) expression was significantly elevated only in grade 3 TNBC non-responders, with no differences in grade 2 TNBC or luminal subtypes. In the prospective cohort, △Copper behavior mirrored this specificity: all grade 3 TNBCs demonstrated positive △Copper, whereas all grade 2 TNBCs showed negative △Copper, and non-responders from other subtypes tended to maintain or reduce systemic copper levels. The only TNBC non-responder, who later relapsed, displayed persistently rising △Copper across the sampling timeline. Taken together, these findings suggest that copper metabolism is tightly linked to both tumor aggressiveness and treatment resistance in high-grade TNBC.

Several biologically plausible mechanisms may explain this pattern. Copper is required for enzymes and signaling mediators such as cytochrome c oxidase, superoxide dismutase, the lysyl oxidase (LOX/LOXL1–4) family, and kinases including MEK1 and ULK1/2. These copper-dependent activities support several processes fundamental to cancer cell survival and progression.^1^ Increased copper import through CTR1 could enhance the activity of these or other copper-dependent pathways, contributing to metabolic fitness, redox stability, autophagy regulation, and ECM remodeling, processes that may collectively promote aggressive tumor behavior and could attenuate chemotherapy efficacy. The LOX family is particularly relevant: LOX-mediated collagen crosslinking has been implicated in chemotherapy resistance in TNBC, and LOX inhibition can re-sensitize resistant models.^21^ Increased copper availability may enhance LOX activity and ECM-mediated survival pathways, providing a plausible mechanism linking copper metabolism to treatment resistance in high-grade TNBC.

We also observed that patients with smaller primary tumors had higher pretreatment serum copper, aligning with the hypothesis that biologically active, early-stage tumors may trigger copper mobilization from copper-rich sources such as the liver to meet angiogenic and metabolic demands. This pattern parallels preclinical findings of elevated intratumoral copper in smaller tumors.^10^ If validated, pretreatment copper levels and △Copper trajectories could serve as stage-or biology-informed biomarkers.

From a translational perspective, combining tumor CTR1 expression with △Copper dynamics emerges as a promising biomarker profile. This integrated copper signature could help identify patients with copper-dependent and treatment-resistant biology, particularly within high-grade TNBC and may guide the selection of patients for copper-modulating therapeutic strategies.

These findings also support the concept that copper biomarkers may complement existing clinicopathologic tools for risk stratification in the neoadjuvant setting.

Although the prospective cohort size is modest, the reproducible, subtype-and grade-specific patterns observed across two independent datasets provide a strong basis for further investigation. Future studies should expand these observations in larger cohorts and incorporate △Copper alongside tumor profiling of CTR1, copper-dependent enzymes, and copper-regulated signaling pathways to refine patient selection and clarify mechanisms linking copper metabolism to neoadjuvant chemotherapy resistance.

In summary, this integrated analysis provides the first clinical evidence that high-grade TNBC activates a coordinated tumor-systemic copper program characterized by elevated CTR1 expression and increased △Copper during neoadjuvant therapy. These hypothesis-generating findings strengthen the emerging link between copper metabolism, tumor aggressiveness, and treatment resistance, and establish a foundation for incorporating copper-related biomarkers into future personalized therapeutic strategies and clinical trial designs.

## 5. Ethics approval and consent to participate

The prospective observational component of this study involving human participants was reviewed and approved by the University of Missouri Institutional Review Board (IRB #2026923). Written informed consent was obtained from all participants prior to enrollment. All clinical samples and associated data were coded, deidentified, and handled in accordance with institutional policies and HIPAA regulations.

The retrospective gene expression analyses were conducted using ROCplot.org, which integrates publicly available, deidentified breast cancer transcriptomic datasets. Analysis of these public datasets did not require additional institutional review board approval and was performed in accordance with the data access and usage policies of the original data sources.

## 6. Data availability

The prospective clinical data supporting the findings of this study are not publicly available due to patient privacy and ethical considerations but may be made available from the corresponding author upon reasonable request and with appropriate institutional approvals. Publicly available gene expression datasets analyzed in this study are accessible through ROCplot.org.

## 8. Author contributions

VCS and CP conceived and designed the study. VCS led data curation, formal analysis, investigation, methodology development, validation, visualization, sample collection and processing, and drafted the original manuscript. CP supervised the study, secured funding, provided resources, contributed to data curation, formal analysis, investigation, methodology development, validation and visualization, and co-wrote the manuscript. NG contributed to data curation, investigation, validation, visualization, and sample collection and processing. MY contributed to data curation and validation. KC contributed to data validation and to sample collection and processing. SAD contributed to sample collection and processing. PS and PR performed formal analyses and contributed to validation and visualization. MJP and LV provided resources and contributed to manuscript review and editing. All authors critically revised the manuscript for important intellectual content and approved the final version.

## Data Availability

All data produced in the present study are available upon reasonable request to the authors.

https://rocplot.com/site/treatment

## 10. Acknowledgments

ICP-MS measurements were performed in the OHSU Elemental Analysis Core with partial support from the National Institutes of Health (S10OD028492). The authors thank Erin Cummings, RN-BSN, OCN, for assistance with blood collection from breast cancer patients.

## References

1. Shanbhag VC, Gudekar N, Jasmer K, et al. Copper metabolism as a unique vulnerability in cancer. Biochim Biophys Acta Mol Cell Res. 2021;1868(2):118893.

2. Zhou B, Gitschier J. hCTR1: a human gene for copper uptake identified by complementation in yeast. Proc Natl Acad Sci U S A. 1997;94(14):7481–6.

3. Nevitt T, Ohrvik H, Thiele DJ. Charting the travels of copper in eukaryotes from yeast to mammals. Biochim Biophys Acta. 2012;1823(9):1580–93.

4. Kuo YM, Zhou B, Cosco D, et al. The copper transporter CTR1 provides an essential function in mammalian embryonic development. Proc Natl Acad Sci U S A. 2001;98(12):6836–41.

5. Nose Y, Kim BE, Thiele DJ. Ctr1 drives intestinal copper absorption and is essential for growth, iron metabolism, and neonatal cardiac function. Cell Metab. 2006;4(3):235–44.

6. Tsang T, Posimo JM, Gudiel AA, et al. Copper is an essential regulator of the autophagic kinases ULK1/2 to drive lung adenocarcinoma. Nat Cell Biol. 2020;22(4):412–24.

7. Brady DC, Crowe MS, Turski ML, et al. Copper is required for oncogenic BRAF signalling and tumorigenesis. Nature. 2014;509(7501):492–6.

8. Rigiracciolo DC, Scarpelli A, Lappano R, et al. Copper activates HIF-1alpha/GPER/VEGF signalling in cancer cells. Oncotarget. 2015;6(33):34158–77.

9. Ramchandani D, Berisa M, Tavarez DA, et al. Copper depletion modulates mitochondrial oxidative phosphorylation to impair triple negative breast cancer metastasis. Nat Commun. 2021;12(1):7311.

10. Ishida S, Andreux P, Poitry-Yamate C, et al. Bioavailable copper modulates oxidative phosphorylation and growth of tumors. Proc Natl Acad Sci U S A. 2013;110(48):19507–12.

11. Glasauer A, Sena LA, Diebold LP, et al. Targeting SOD1 reduces experimental non-small-cell lung cancer. J Clin Invest. 2014;124(1):117–28.

12. Sajesh BV, McManus KJ. Targeting SOD1 induces synthetic lethal killing in BLM-and CHEK2-deficient colorectal cancer cells. Oncotarget. 2015;6(29):27907–22.

13. Shanbhag V, Jasmer-McDonald K, Zhu S, et al. ATP7A delivers copper to the lysyl oxidase family of enzymes and promotes tumorigenesis and metastasis. Proc Natl Acad Sci U S A. 2019;116(14):6836–41.

14. Cox TR, Bird D, Baker AM, et al. LOX-mediated collagen crosslinking is responsible for fibrosis-enhanced metastasis. Cancer Res. 2013;73(6):1721–32.

15. Jasmer KJ, Shanbhag VC, Forti KM, et al. Pulmonary lysyl oxidase expression and its role in seeding Lewis lung carcinoma cells. Clin Exp Metastasis. 2024;42(1):7.

16. Chan N, Willis A, Kornhauser N, et al. Influencing the Tumor Microenvironment: A Phase II Study of Copper Depletion Using Tetrathiomolybdate in Patients with Breast Cancer at High Risk for Recurrence and in Preclinical Models of Lung Metastases. Clin Cancer Res. 2017;23(3):666–76.

17. Fekete JT, Gyorffy B. ROCplot.org: Validating predictive biomarkers of chemotherapy/hormonal therapy/anti-HER2 therapy using transcriptomic data of 3,104 breast cancer patients. Int J Cancer. 2019;145(11):3140–51.

18. Favier A, Ruffieux D. Physiological variations of serum levels of copper, zinc, iron and manganese. Biomed Pharmacother. 1983;37(9-10):462–6.

19. McMaster D, McCrum E, Patterson CC, et al. Serum copper and zinc in random samples of the population of Northern Ireland. Am J Clin Nutr. 1992;56(2):440–6.

20. Taylor AA, Tsuji JS, Garry MR, et al. Correction to: Critical Review of Exposure and Effects: Implications for Setting Regulatory Health Criteria for Ingested Copper. Environ Manage. 2022;69(5):1051.

21. Saatci O, Kaymak A, Raza U, et al. Targeting lysyl oxidase (LOX) overcomes chemotherapy resistance in triple negative breast cancer. Nat Commun. 2020;11(1):2416.

